# Transcriptome analysis of PBMCs reveals distinct immune response in the asymptomatic and re-detectable positive COVID-19 patients

**DOI:** 10.1101/2021.03.16.21251286

**Authors:** Jiaqi Zhang, Dongzi Lin, Kui Li, Xiangming Ding, Lin Li, Yuntao Liu, Dongdong Liu, Jing Lin, Xiangyun Teng, Yizhe Li, Ming Liu, Xiaodan Wang, Dan He, Yaling Shi, Dawei Wang, Jianhua Xu

## Abstract

The existence of asymptomatic and re-detectable positive COVID-19 patients presents the disease control challenges of COVID-19. Most studies on immune response of COVID-19 have focused on the moderately or severely symptomatic patients, however little is known about the immune response in asymptomatic and re-detectable positive patients. Here we performed a comprehensive analysis of the transcriptomic profiles of PBMCs from 48 COVID-19 patients which include 8 asymptomatic, 13 symptomatic, 15 recovering and 12 RP patients. Our analysis revealed a down-regulation of IFN response and complement activation in the asymptomatic patients compared with the symptomatic, indicating a weaker immune response of the PBMCs in the asymptomatic patients. In addition, we observed a lower expression of the cytokines and chemokines in the PBMC of asymptomatic and symptomatic patients. In contrast, the cytokines and chemokines level in the RP patients are higher than the recovering. GSEA analysis showed the enrichment of TNFa/NF-κB and influenza infection in the RP patients compared with the recovering patients, indicating a flu-like, hyper-inflammatory immune response in the PBMC of RP patients. Thus our findings could extend our understanding of host immune response during the progression COVID-19 disease and help the clinical management and the immunotherapy development for COVID-19.

## Introduction

Coronavirus disease 2019 (COVID-19) is an infectious disease caused by severe acute respiratory syndrome coronavirus 2 (SARS-CoV-2), which was declared a pandemic by the WHO on 11 March 2020[1]. As of 14 November 2020, 53 million cases have been reported across 188 countries and territories with more than 1.29 million deaths. Most COVID-19 patients were classified as mild; however, about 20% become seriously ill and require hospitalization due to pneumonia [2]. The primary symptoms of COVID-19 are listed as fever, dry cough, and shortness of breath but also include other symptoms such as diarrhea, loss of taste and smell, and rashes [3]. However, increasing evidence has shown that individuals infected with SARS-CoV-2 can be asymptomatic and simultaneously silent spreaders of SARS-CoV-2 [4–5]. Identification and isolation of asymptomatic patients as early as possible is critical to control COVID-19 outbreaks. According to a recent study, four medical workers aged from 30 to 36 years who had re-detectable positive (RP) for SARS-CoV-2 within 5–13 days after recovery, suggesting that these four recovered patients may still be virus carriers [6]. All these four patients continued to be asymptomatic by clinician examination and chest CT findings showed no change from previous images. RP cases have also been reported by several other studies. For example, one report showed that 10.99% of patients (20/182) detected SARS-CoV-2 RNA re-positive and none of them showed any clinical symptomatic recurrence [7]. Now both the asymptomatic patients and RP patients are receiving more attention due to the potential infectivity of these patients.

As there are no effective drugs or vaccines available at this moment against SARS-CoV-2 and most patients are receiving symptomatic treatment, it is essential to understand the host– pathogen biology of COVID-19 which will provide the novel insights into more targeted therapeutic approaches of this disease. Over the past few months, studies have reported the characteristics of innate and adaptive immune responses of SARS-CoV-2 infection, which have helped us to understand the potential pathogenesis of COVID-19[8–9]. Most of these researches focused on the peripheral immune response of the symptomatic patients, particularly the severe COVID-19 patients [10–11]. For the severe COVID-19 patients, a dysfunctional immune response was observed, which triggers a cytokine storm that cause severe lung and even systemic pathology [12]. However, the immunological features and the molecular mechanisms involved in the asymptomatic and RP patients still remains elusive. Here we report the transcriptome profiles of PBMCs from COVID-19 patients composed of asymptomatic, symptomatic, recovering and RP groups. Our study revealed a weaker peripheral immune response in the asymptomatic group and a hyper-inflammatory, flu-like immune response in the RP group.

## Materials and methods

### Patients

This study was reviewed and approved by the ethics committees of the above four participant hospitals. Written informed consent was obtained from all participants enrolled in this study. From March and May in 2020, 48 patients with confirmed SARS-CoV-2 admitted to Guangzhou Eighth People’s Hospital, Shunde Hospital of Guangzhou University of Chinese Medicine, Fourth People’s Hospital of Foshan, and First People’s Hospital of Foshan, were enrolled in this study, including four groups. Patients in the Asymptomatic group (8 cases) without self-perceived or clinically recognizable symptoms but diagnosed as having COVID-19 according to the Protocol for Prevention and Control of COVID-19 (Edition 6) of the National Health Commission of China. According to the Guidelines for the Diagnosis and Treatment of COVID-19 (7th Edition), symptomatic group (13 cases) were defined as patients with evident clinical symptoms, and convalescent group (15 cases) were defined as patients met the discharge standards. The re-detectable positive group (12 cases) were those who with reLJpositive results of SARS-CoV-2 nucleic acid during the follow-up period after discharge from hospital. In addition, we had enrolled 22 healthy donors from Shunde Hospital of Guangzhou University of Chinese Medicine. All patients had routine laboratory investigations, including complete blood count; liver function tests, blood gases analysis, and coagulation tests. The PBMC samples were collected within 4 days of admission from asymptomatic and symptomatic groups to maintain uniformity of timing for comparison between groups.

### RNA extraction and library construction

Total RNA was isolated and purified using TRIzol(Life, cat.265709, CA, USA) following the manufacturer’s procedure. After the quality inspection of Agilent 2100 Bioanalyzer (Agilent, cat.G2939AA, CA, USA) and NanoPhotometer® (Implen, cat.N60, Munich, Germany), poly (A) RNA is purified from 1μg total RNA using VAHTS® mRNA Capture Beads with Oligo (dT) (Vazyme, cat.N401-01, Nanjing, China) using two rounds of purification. Then the poly(A) RNA was fragmented into small pieces using VAHTS® Universal V6 RNA-seq Library Prep Kit (Vazyme, cat.NR604, Nanjing, China) under 94□ 8min. Then the cleaved RNA fragments were reverse-transcribed to create the cDNA by reverse transcription reagent, which were next used to synthesise U-labeled second-stranded DNAs. An A-base was then added to the blunt ends of each strand, preparing them for ligation to the indexed adapters. Each adapter contains a T-base overhang for ligating the adapter to the A-tailed fragmented DNA. After the heat-labile UDG enzyme treatment of the U-labeled second-stranded DNAs, size selection was performed with VAHTS® DNA Clean Beads (Vazyme, cat. N411, Nanjing, China). Then the ligated products are amplified with PCR. At last, we performed the 2×150bp paired-end sequencing (PE150) on an Illumina Novaseq™ 6000 (Guangzhou Huayin Health Technology Co., Ltd. Guangzhou, China) following the vendor’s recommended protocol.

### Data processing and analysis

Cutadapt software (cutadapt v1.16) was used to remove the reads contained adaptor contamination. Then we got the CleanData after removing the low quality bases and undetermined bases. The clean reads were mapped to the hg19 UCSC transcript set using Bowtie2 version 2.1.0 and the gene expression level was estimated using RSEM v1.2.15. 24 immunologically relevant gene sets were collected from ImmPort database and molecular signature database. The detailed lists of the gene sets were in the supplementary table 1. Gene sets were scored using single-sample gene set enrichment (ssGSEA) analysis, as implemented in the GSVA R package. The deconvolution analysis was performed using the MCP-counter method to calculate the abundance of each immune cell in PBMC of the COVID-19 patient and healthy donors. The statistical analyses were performed using a two-tailed t-test. All statistical analyses were performed using R software (version 4.0.2).

## Results

### Differences in immune cells abundance across disease conditions

To investigate the immunological features of asymptomatic and RP patients with COVID-19, we performed the mRNA sequencing to study the transcriptomic profiles of PBMCs from 48 COVID-19 patients which include 8 asymptomatic, 13 symptomatic, 15 recovering and 12 RP patients. 22 healthy donors were also included as the control group. The demographic, clinical and laboratory characteristics of enrolled patients are summarized in Table 1.The laboratory result revealed a significant lower level of lymphocytes/leukocytes in symptomatic patients compared with the healthy donors (Fig.1A), which is consistent with previous report of lymphocytopenia/leukopenia in COVID-19 patients[2]. In addition, eosinophils and basophils counts are also significantly lower than in symptomatic patients compared with the healthy donors, although no significant difference were identified for the monocytes and neutrophils counts (Fig.1A). As shown in Fig.1A, the lymphocytes counts in asymptomatic patients are higher than in symptomatic patients, although the difference is not significant due to the small sample size. The basophils counts in asymptomatic patients are significant higher compared with the symptomatic patients. Interestingly, a significant lower level of monocytes in RP patients was observed when compared with the recovering patients and it is comparable for other blood cell counts between RP and recovering patients.

**Table1.** Baseline characteristics and laboratory parameters of patients infected with SARS-CoV-2.

|  | Healthy<br>( n=22 ) | Asymptomatic<br>( n=8 ) | Symptomatic<br>( n=13 ) | Recovering<br>( n=15 ) | Re-detectable<br>positive ( n=12 ) |
| --- | --- | --- | --- | --- | --- |
| Age (y, median and IQR) | 30.50(26.75-39.00) | 28.50(21.50-40.75) | 33.00(26.00-48.50) | 56.00(25.00-68.00) | 35.00(25.25-59.75) |
| Female (n, %) | 5(22.7%) | 4(50%) | 11(84.6%) | 5(33.3%) | 6(50%) |
| Male (n, %) | 17(77.3%) | 4(50%) | 2(15.4%) | 10(66.7%) | 6(50%) |
| Signs and symptoms (n, %) |  |  |  |  |  |
| Cough |  |  | 5(38.5%) | 3(20.0%) | 1(8.3%) |
| Expectoration | 0 | 0 | 5(38.5%) | 3(20.0%) | 1(8.3%) |
| Shortness of breath | 0 | 0 | 4(30.8%) | 1(6.7%) | 1(8.3%) |
| Fatigue | 0 | 0 | 4(30.8%) | 0(0) | 1(8.3%) |
| Sore throat | 0 | 0 | 3(23.1%) | 0(0) | 1(8.3%) |
| Chest tightness | 0 | 0 | 3(23.1%) | 1(6.7%) | 0(0) |
| Dry throat | 0 | 0 | 3(23.1%) | 0(0) | 1(8.3%) |
| Headache | 0 | 0 | 3(23.1%) | 0(0) | 0(0) |
| Chills | 0 | 0 | 0(0) | 1(6.7%) | 1(8.3%) |
| Poor stomach intake | 0 | 0 | 1(7.7%) | 0(0) | 1(8.3%) |
| Gastrointestinal reaction | 0 | 0 | 0(0) | 0(0) | 1(8.3%) |
| Palpitation | 0 | 0 | 0(0) | 0(0) | 1(8.3%) |
| Diarrhea | 0 | 0 | 0(0) | 1(6.7%) | 0(0) |
| Myalgia | 0 | 0 | 0(0) | 1(6.7%) | 0(0) |
| Comorbidities (n, %) |  |  |  |  |  |
| Hypertension | 0 | 0 | 2(15.4%) | 1(6.7%) | 4(33.3%) |
| Diabetes | 0 | 0 | 2(15.4%) | 1(6.7%) | 2(16.7%) |
| Liver disease | 0 | 0 | 1(7.7%) | 1(6.7%) | 1(8.3%) |
| digestive diseases | 0 | 0 | 1(7.7%) | 1(6.7%) | 0 |
| Kidney disease | 0 | 0 | 0 | 1(6.7%) | 1(8.3%) |
| Anemia | 0 | 0 | 0 | 1(6.7%) | 1(8.3%) |
| Cardiovascular disease | 0 | 0 | 0 | 1(6.7%) | 0 |
| Thyroid disease | 0 | 1(12.5%) | 0 | 0 | 0 |
| Hypertriglyceridemia | 0 | 1(12.5%) | 0 | 0 | 0 |
| Rhinitis | 0 | 0 | 1(7.7%) | 0 | 0 |
| laboratory parameters ( median and IQR) |  |  |  |  |  |
| Leukocytes ($\times 10^9/L$) | 6.71(6.05-7.83) | 6.19(5.16-7.29) | 4.68(4.11-6.74) | 5.20(4.87-7.86) | 5.80(4.65-7.71) |
| Lymphocytes ($\times 10^9/L$) | 2.21(1.79-2.46) | 1.71(1.35-2.51) | 1.29(0.92-1.94) | 1.78(1.51-2.18) | 1.69(1.23-2.32) |
| Monocytes ($\times 10^9/L$) | 0.40(0.31-0.49) | 0.36(0.27-0.48) | 0.35(0.25-0.47) | 0.38(0.34-0.57) | 0.29(0.23-0.34) |
| Neutrophils ($\times 10^9/L$) | 3.95(3.51-4.40) | 3.44(2.89-3.98) | 2.75(2.32-4.28) | 3.02(2.19-5.30) | 3.67(2.72-4.85) |
| Eosinophils ($\times 10^9/L$) | 0.14(0.10-0.33) | 0.06(0.04-0.18) | 0.01(0.01-0.13) | 0.12(0.08-0.30) | 0.10(0.06-0.18) |
| Basophils ($\times 10^9/L$) | 0.04(0.02-0.05) | 0.03(0.02-0.05) | 0.02(0.01-0.03) | 0.02(0.01-0.02) | 0.02(0.01-0.03) |

**Figure 1.**
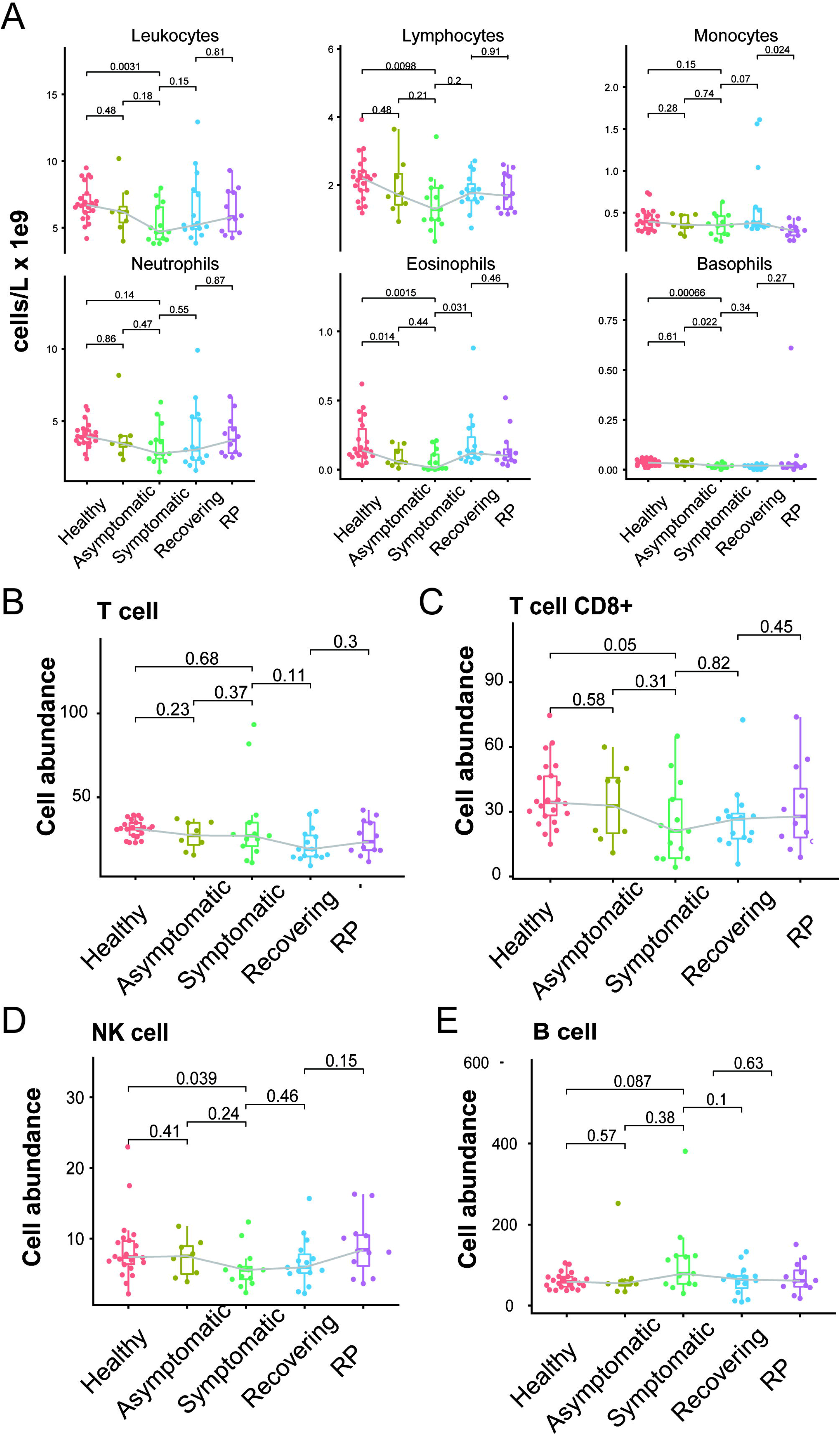
Comparison of immune cell abundance among the COVID-19 patients and healthy donors. A) Clinical complete blood counts with differential cell count for COVID-19 patients and healthy donors. B-E) Cell abundance of subset of the lymphocytes calculated by MCPcounter method in COVID-19 patients and healthy donors. Each dot represents the value of each individual from the groups (healthy donors, n = 22; asymptomatic, n = 8; symptomatic, n = 13; recovering, n= 15; RP, n=12). The box plots show the medians (middle line) and first and third quartiles (boxes), and the whiskers show 1.5× the IQR above and below the box. Unpaired, two-sided T test p values are depicted in the plots.

To characterize the changes of subset cells of the lymphocyte across these groups, we performed a deconvolution analysis [13] to detect the abundance of each immune cell in each patients based on the RNAseq data of the PBMC. T and NK cells play critical roles in viral clearance during respiratory infections [13]. As shown in Fig.1C and D, the abundance of CD8+ T and NK cells are significantly lower in symptomatic patients compared with the healthy donors, which is consistent with previous reports [14–15]. However, the abundance of CD8+ T and NK cells are comparable in symptomatic patients and healthy donors. We did not observe any significant difference of total T and B cell abundance among these groups.

### Differences in immune responses across disease conditions

To identify the difference of immune-related function among these 5 groups, we collect 24 immunologically relevant gene sets including 17 from ImmPort database [16] and 7 from the Hallmark gene set in molecular signature database [17]. These 24 immune-associated gene sets represented diverse immune functions and pathways (Supplementary Table 1). We used the ssGSEA score [18–19] to quantify the activity or enrichment levels of immune-related functions or pathways in these PBMCs samples. Multiple immune-related functions were dysregulated in PBMCs from COVID-19 patients. The ssGSEA analysis revealed a dynamic change of the interferon response among these five groups. As expected, both the interferon_alpha_response and interferon_gamma_response activity are significantly higher in the symptomatic group compared with the healthy donors (Fig.2A and B). Interestingly, we observed that the levels of interferon response of asymptomatic group are comparable with the healthy group and significantly lower than the symptomatic group, indicating a lower level of IFN in the serum of asymptomatic patients. NK cells are an important arm of the innate lymphocytic antiviral response. The activity of NK cell cytotoxicity is significantly lower in both asymptomatic and symptomatic when compared with the healthy group (Fig.2E). The RP patients have a significant higher level of NK cell cytotoxicity than the recovering patients, indicating a restoration of the dysregulated immune system. In addition, both the symptomatic and asymptomatic patients have a significant lower level of antigen processing and presentation than the healthy donors, suggesting a defective antigen presentation in the COVID-19 patients (Fig.2C). Interestingly, we also observed the RP patients have a restored antigen presentation to the normal level. Furthermore, the level of cytokines, chemokines, and interleukins in the PBMCs showed a similar pattern as the antigen processing and presentation (Fig.2D and Supplementary Fig.1). The lower expression of cytokines in symptomatic patients is consistent with previous reports of undetectable expression of most cytokines in the PBMCs of COVID-19 patients [8, 20]. The lower level of cytokines in PBMC indicates the elevated serum cytokines largely arise from the local infection site instead of the peripheral blood. On the contrary, the levels of cytokines, chemokines, and interleukins in the RP group are significantly higher than in other groups of COVID-19 patients, indicating a hyper-inflammatory immune response in the PBMC of RP patients.

**Figure 2.**
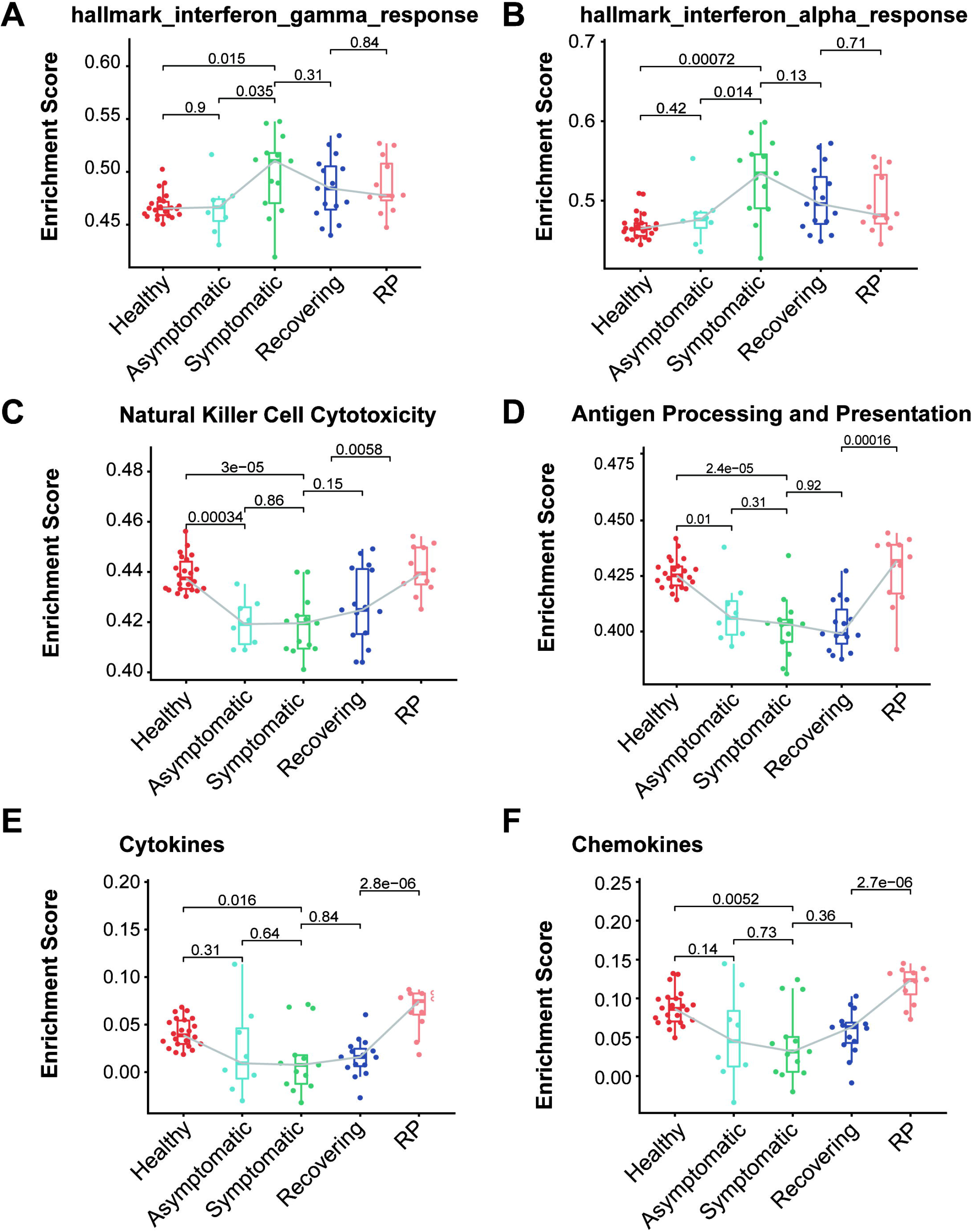
Comparison of immune-relate genesets activity among the COVID-19 patients and healthy donors. A-F) Boxplot of ssGSEA enrichment scores of hallmark_interferon_gamma_response(A),hallmark_interferon_alpha_response(B), Natural Killer Cell Cytotoxicity(C), Antigen Processing and Presentation (D), Cytokines(E) and Chemokines(F) in individuals from each group (healthy donors, n = 22; asymptomatic, n = 8; symptomatic, n = 13; recovering, n= 15; RP, n=12). The box plots show the medians (middle line) and first and third quartiles (boxes), and the whiskers show 1.5× the IQR above and below the box. Unpaired, two-sided T test p values are depicted in the plots, and the significant p value cutoff was set at 0.05.

To further elucidate the up- and down-regulated biological pathways in asymptomatic verse symptomatic and RP verse recovering groups, we performed the GSEA analysis [21] to interrogate the pathways from KEGG, Reactome and Biocarta as well as the hallmark genesets from MSigDB. We observed the down-regulation of IFN response and complement activation (“Creation of C4 and C2 activators”) in the asymptomatic patients (Fig.3 A and C), indicating a weaker immune response of the PBMCs in the asymptomatic patients. Interestingly, some cell cycles related pathways were down-regulated and NGF related pathway up-regulated in the asymptomatic patients. The potential role of these pathways needs further investigation. In addition, both the TNFa/NF-κB and influenza infection activity were enriched in the RP patients compared with the recovering patients (Fig.2B and D). NF-κB is one of the hallmark signaling factors activated by Influenza infection[22]. Moreover, a comparative study of COVID-19 and influenza showed the activation of STAT3/NF-κB in PBMCs of influenza A virus (IAV)-infected patients vs the activation of STAT1/IRF3 in COVID-19 patients. Taken together, these findings suggest the RP patients have a flu-like immune response in the PBMCs.

**Figure 3.**
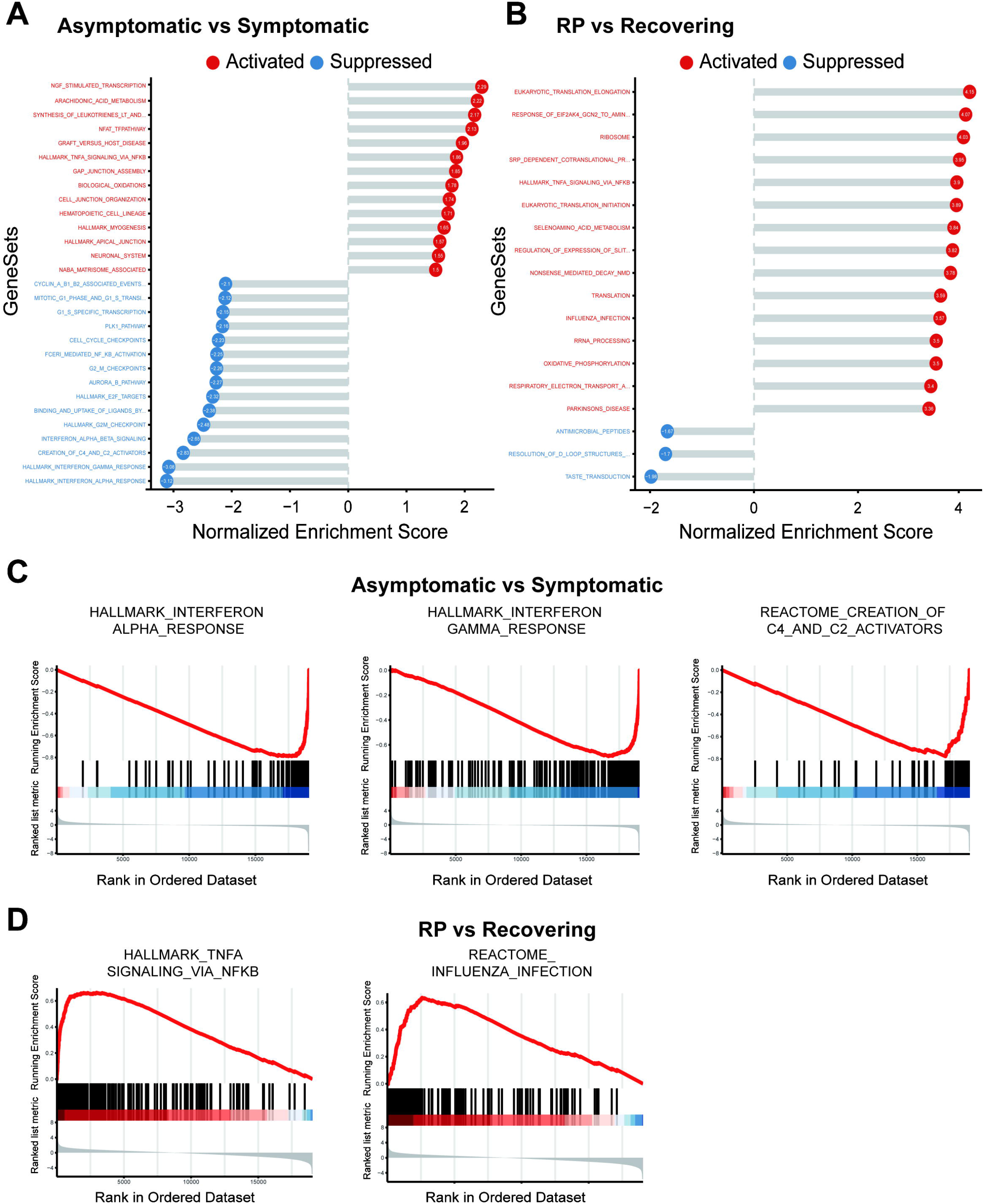
GSEA analysis of the comparison of asymptomatic verse symptomatic and RP verse recovering groups. A-B) The top significantly up- and down-enriched genesets in the comparison of asymptomatic verse symptomatic (A) and RP verse recovering groups(B). The status of the genesets are colored by the red (activation) and blue (suppression). C) Curves of GSEA enrichment scores for the IFN response and complement activators in asymptomatic verse symptomatic. D) Curves of GSEA enrichment scores for the NF-κB signaling and Influenza infection in RP verse recovering.

## Discussion

COVID-19 patients are characterized by a broad spectrum of disease severity ranging from asymptomatic to critically severe [23]. The heterogeneous manifestation of the disease may be due to the different immune response to SARS-CoV-2 by the patients. Dysregulated immune responses have been described in symptomatic COVID-19 patients, particularly of severe disease [10, 24]. However, little is known about the immune response in the asymptomatic and re-detectable positive patients.

Here, we performed the transcriptome analysis on PBMCs from 48 COVID-19 patients in different phases including the asymptomatic and RP patients, as well as 22 healthy donors. We found the level of IFN response and complement activation in asymptomatic patients is lower than in the symptomatic patients. This observation is agreement with the data showing lower levels of 18 cytokines including the IFN in the serum of the asymptomatic compared with the symptomatic patients. It was reported that plasma IFN levels was associated with the COVID-19 disease severity [25] and a recent longitudinal analysis showed that IFNα in peripheral blood was sustained at high levels in patients with severe COVID-19 [26]. A robust type I interferon response could exacerbate hyper-inflammation in the progression to severe COVID-19 through diverse mechanisms [27]. In addition, two recent studies of serological responses in COVID-19 patients revealed that the asymptomatic patients did not respond or have lower antibody levels upon SARS-CoV-2 infection [28–29]. Moreover, the asymptomatic patients were reported to exhibit reduced proportions of SARS-CoV-2 specific T cells, suggesting a weaker cell-mediated immune response [29]. Collectively, the well-controlled immune response in the asymptomatic patients may protect the patients from progressing into the inflammatory secondary phase of the disease.

Several recent studies have reported the existence of RP patients and the underlying mechanism of RP occurrence remains unknown [6, 30]. The potential reasons might be related to some factors including virology, immunology and sampling methodology. Factors such as false negatives of the detection [31], viral residual [32], intermittent viral release [33] and viral distribution [34] are usually considered as major reason. However, to the best of our knowledge, there is no report of the peripheral immune response of the RP patients. In this study, we identified a flu-like, hyper-inflammatory immune response in the peripheral blood of RP patients.

Several limitations should be taken into consideration for the interpretation of this study. First, this study focused on the transcriptome analysis of PBMCs in blood. Adding the data of immune cells from lesion sites such as the lung and bronchoalveolar lavage fluid will make the analysis more comprehensive and conclusive. Second, the PBMC samples were collected within 4 days of admission from asymptomatic and symptomatic groups to maintain uniformity of timing for comparison between groups. Future studies with longitudinal samples from COVID-19 patients may help to determine the cause-and-effect relationships between immune response and disease outcome.

In summary, this study will help us extend our understanding of host immune response during the progression COVID-19 disease, and may help elucidate the COVID-19 infection and provide a basis for rationally designed immune therapies.

## Data Availability

Data sharing is not applicable to this article as no new data were created or analyzed in this study.

## Acknowledgements

This study was supported by the Science and Technology Innovation Project of Foshan Municipality (2020001000431) and the National Key Research and Development Project (2020YFA0708001).

## Figure legends

**Supplementary Fig.1.**
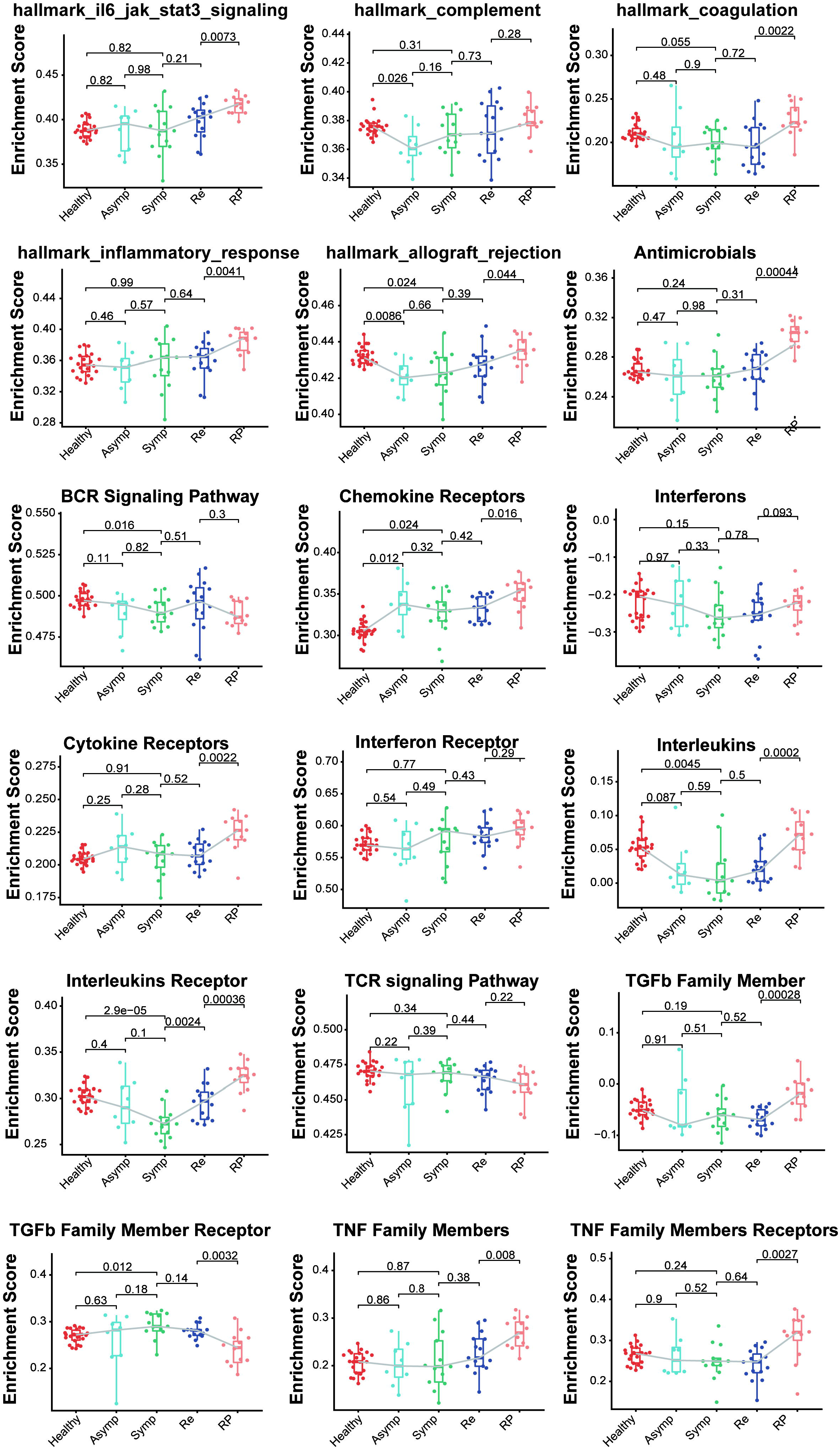
Comparison of immune-relate genesets activity among the COVID-19 patients and healthy donors.

